# Population-based serosurveys for SARS-CoV-2 transmission 2021-2022, Massachusetts USA

**DOI:** 10.1101/2025.01.17.25320194

**Authors:** Estee Y. Cramer, Augustine O. Dada, Shauna Onofrey, Jessica Pearlman, John Cluverius, Rebecca Loveland, Ashley Eaton, Megan Hatch, Lily Harris, Joshua Dyck, Mark Melnik, Peter Reinhard, Andrew A. Lover, Monina R. Klevens

## Abstract

**Background:** SARS-CoV-2 has been responsible for extensive morbidity and mortality in Massachusetts, especially from 2021 – 2022. The true burden of infection is unknown as official reporting data during 2021 and 2022 was not able to capture subclinical/asymptomatic infections nor the results from home-based lateral flow tests (LFTs).

**Aim:** This study was designed to better characterize the exposure of Massachusetts residents to SARS-CoV-2, and to understand demographic and behavioral factors associated with SARS-CoV-2 exposure during the highest burden years of the pandemic.

**Methods:** A series of five sequential state-wide serosurveys were conducted with oversampling for underrepresented demographic groups from June 2021 to September 2022. These mail-based, repeated cross-sectional surveys (RCSs) captured data at periods of rapid vaccine uptake when different viral variants were predominant. This study also included collection of at-home nasal swabs for PCR-based SARS-CoV-2 virological testing, and collection of dried blood spot cards for ELISA-based testing of SARS-CoV-2 IgG antibody markers including spike and capsid, reported as seroprevelences. Neutralizing antibodies to spike-RBD were also measured.

**Results:** Of the randomly selected 249,000 Massachusetts households invited to participate in this survey, a total of 2,220 participants completed the demographic questionnaire and submitted valid specimens for analysis. Of these participants, ten were PCR-positive for SARS-CoV-2 at time of survey. Across all five repeated cross-sectional surveys (RCS), spike antibody positivity ranged from 83.1% to 96.4%. Additionally, levels of the spike neutralizing antibody increased with each RCS; point prevalence values ranged from 20.5% in RCS 1 and 2 to 73.5% in RCS 5. Using weighted data, the seroprevalence of capsid remained relatively constant throughout the RCSs except for RCS 3. Multivariable regression results found a positive association between vaccination status and markers of SARS-CoV-2 exposure; individuals who had been vaccinated were more likely to be seropositive for all markers. Factors including race, age, income, and occupation did not show any statistically significant associations with serostatus.

**Conclusions:** This survey indicates that while there was an increase in antibodies to spike protein and to associated neutralizing antibodies over time, there were no differences in neutralizing antibodies by socio-demographic factors, suggesting no major health disparities existed at the time of surveys in either vaccine coverage or infection-related antibody titers across the state. Response rates were higher among some demographic groups (Caucasians, households with a high income, and women), thus, oversampling and weighting allowed estimates of the larger Massachusetts population. Our findings that there were no statistically significant differences in neutralizing antibodies across demographic groups suggesting all groups were similarly protected from SARS-CoV-2 infection. These results highlight the success of Massachusetts in protecting individuals across all demographics.

## I. Introduction

SARS-CoV-2 has led to extensive morbidity and mortality in the United States including Massachusetts (1–3). By June 2021, there were over 663,000 reported cases of COVID-19 among Massachusetts residents, representing approximately 10% of the Massachusetts population (4). The primary data sources on disease burden during this stage of the pandemic were clinical testing, hospitalizations, and mortality reporting to the Massachusetts Department of Public Health (DPH). These data sources were largely unable to capture sub-clinical/asymptomatic infections, or the results from lateral flow test (LFT) self-testing.

To capture asymptomatic infections, serological testing can be used to measure SARS-CoV-2-specific antibodies. Surveys and serosurveys can be conducted to estimate population-level seropositivity, and understand risk factors associated with infection (5–7). With the application of a rigorous population sample frame and repeated cross-sectional surveys, robust estimates of population-level immunity can be estimated (8).

Prior serosurveys on SARS-CoV-2 antibodies utilized convenience samples or were focused on specific geographies or populations (e.g. blood donors) (9). These samples may not have provided fully representative estimates across the state’s 6.9 million residents (9–12). Moreover, as home tests (LFTs) became available between the 2021 Delta and Omicron waves, routine health sector data became increasingly less representative of underlying transmission dynamics (13).

We conducted a series of five population-based cross-sectional serosurveys as part of the “Get Back Massachusetts” program, from June 2021 - September 2022. These surveys captured data during periods of rapid vaccine uptake and when different viral variants were predominant. These mail-based surveys used at-home bio-sample collection for PCR-based virological testing and ELISA-based serology, with oversampling for underrepresented groups for RCSs 3-5. The study objectives were to 1) measure the population prevalence of antibodies by selected characteristics during 2021-2022, and 2) identify exposures associated with vaccine- and infection-related immunity.

## II. Methods

### Study design

We conducted a series of five repeated cross-sectional surveys (RCS) from June 2021 through September 2022 during the following time periods: RCS 1: June 2-July 11, 2021; RCS 2: July 29-October 16, 2021; RCS 3: October 20, 2021 - January 4, 2022; RCS 4: January 10 - May 5, 2022; RCS 5: June 1 - September 11, 2022). For data collection, a “push to the web” approach was used. Postcards with QR codes and URLs were mailed to homes in Massachusetts, and participants were guided to an online portal where they could answer a series of survey questions. After informed consent, participants were mailed a bio-sample collection kit (Supplemental Figure 1), which included detailed instructions on how to collect and return biospecimens for laboratory analysis.

### Participant recruitment

The target population included all Massachusetts residents aged 18 or older at the time of the survey. Data collection extended over multiple waves of variants beginning with Delta, capturing the large first Omicron wave, and ending with the subsequent Omicron sub-lineages (Figure 1).

**Figure 1:**
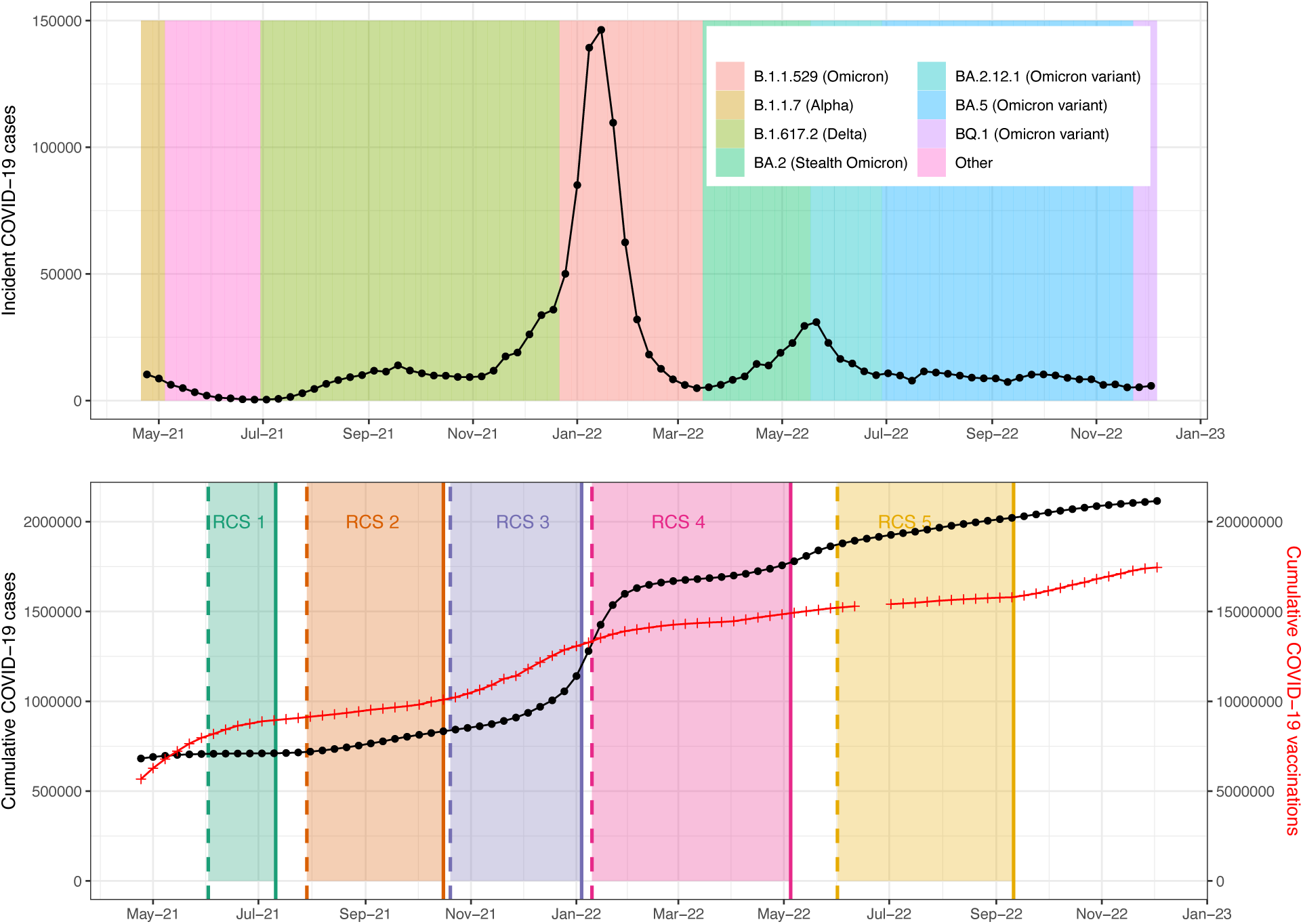
Overview of study repeated cross-sectional surveys (RCSs), state-level vaccine uptake, and predominant viral variants, “Get Back Massachusetts” study, June 2021 - September 2022. Notes: Data extracted from covariants.org, and from https://www.mass.gov/info-details/massachusetts-covid-19-vaccination-data.

To recruit participants, a sample of Massachusetts residents was generated for this study by Data Axle – a data source that provides consumer records to a variety of commercial, nonprofit, and research-based clients (14). This source was chosen due to reported high accuracy of mailing addresses, as well as inclusion of detailed education and ethnicity data to ensure representative samples.

Each RCS was initiated with three discrete postcard distributions, sent one week apart to each address in the sample frame. Repeated mailings were used to increase participation rates. From each address, only one individual was eligible to participate in the study. Participants who completed the demographic survey were sent a $25 gift card for an online retailer and were also entered into a drawing for a $1,000 gift card. Finally, participants were also sent their study results.

### Oversampling demographic groups

For RCS 1, postcards were mailed to 4,000 households; RCS 2 was expanded to include 16,000 selected households. Preliminary analyses after RCS 2 identified lower-than-expected response rates among sampled Black households. Consequently, RCS 3 had a sample size of 16,000 households, which included an oversample of 3,000 households with at least one Black individual. Similarly, we identified lower-than-expected response rates among households with lower levels of educational attainment, therefore in RCSs 4 and 5, the sample size of 16,000 households included an oversample of 3,000 households with at least one Black individual, and 3,000 households with at least one adult (age 18 or higher) without a high school diploma.

### Survey design

The survey questions included items related to socio-demographic characteristics and identification as an essential worker. Vaccine status was also collected as a self-report through this survey. Self-reported vaccine data was cross-validated with data from state vaccination records at DPH. Across the RCS round, data were stored in Qualtrics (15), Salesforce (16), and REDCap (17) due to changing data management needs across the study period. Final datasets were compiled in a HIPAA-compliant environment (University-administered virtual machines).

### Biospecimen Collection and Laboratory Analysis

Upon completion of the online survey, participants were mailed test kits with materials for self-collection of dried blood spots (DBS) and nasal samples with detailed instructions for finger-stick and nasal collection. Nasal swabs were tested for SARS-CoV-2 via Reverse Transcriptase Quantitative PCR (RT-qPCR).

For serology, dried blood spot (DBS) samples were extracted and tested for markers using an ELISA-based assay to detect the anti-Receptor Binding Domain (spike anti-RBD). Samples testing positive for anti-RBD (values above background-corrected detection threshold, OD_405_ = 0.36) were assigned as positive. A traditional ELISA was also used for nucleocapsid protein (capsid), a traditional ELISA was conducted using the DBS. Cut-off values for capsid IgG results were established through threshold optimization experiments with known positive and negative samples.

All DBS samples that were positive for anti-RBD antibodies were subsequently tested for neutralizing antibodies using a cell-based neutralization antibody assay. This neutralizing antibody measurement is a measurement of recent antigen exposure, either as a COVID-19 vaccination or due to natural infection. Samples that passed the assay quality control and showed neutralizing efficiency value ≥ 50% were categorized as positive for SARS-CoV-2 neutralizing antibodies.

### Statistical analysis and analytical plan

To adjust for the complex sample frame, population-level prevalences were adjusted for underreporting to generate a sample more representative of the adult Massachusetts population using iterative proportional fitting (IPF) raking, standardized to the American Community Survey (ACS). This was done by implemented separately for each RCS. A subset of the ACS data curated by the Integrated Public Use Microdata Series (IPUMS) from the years 2017 through 2021 (18) was used. Weighting used the R package, *PSweight* (19) using the variables of age, race, sex, and educational attainment. No imputation was done; complete case records were used for raking weights. Due to the limited number of respondents in RCS 1, this group was aggregated with RCS 2 for analyses.

To assess factors associated with capsid seropositivity (an indicator of infection) and neutralizing antibodies (an indicator of vaccination and/or past infection) multivariable models were developed. These models used Poisson regression with robust standard errors and exponentiated coefficients, reporting estimates as prevalence ratios (PRs) (20). As a sensitivity analysis for capsid antibodies, the untransformed titer data (as opposed to a binary seropositivity) were also modeled using a multivariable quantile regression for the median values. (21).

For all models, variables were selected that had a p-value below 0.2 or have been reported in the literature to be associated with SARS-CoV-2 infection. Sex and race/ethnicity were also forced into these models. Akaike/Bayesian Information Criteria (AIC/BIC) were used to build parsimonious models.

To compare capsid IgG antibody titers between vaccinated and unvaccinated individuals across the RCS rounds, a Kruskal-Wallis test was used. Pairwise comparisons were conducted using Dunn’s test, with Bonferroni correction applied to adjust for multiple comparisons. Analyses were performed using R version 4.1.2 (22). All tests were two-tailed, with alpha = 0.05.

### Ethical statement

The Get Back Massachusetts survey was approved by the UMass IRB (Approval #2664; date, April 8, 2021). Written informed consent was obtained from all participants.

## III. Results

### Survey response

Of all residents invited to participate in this survey, a total of 3,717 (1.6%) responded, were eligible to participate, and were sent biospecimen test kits. Of these, 2,302 participants (86.0%) returned the kits. A small number of samples did not meet quality cutoffs and were excluded. In total, 2,220 individuals submitted valid survey data and valid biospecimens (Supplemental Figure 4); demographics of these participants are shown in Table 1. Of the 2,220 participants included in this study, 1,948 participants (87.7%) had complete case data for all variables of interest; this population was used to estimate the weighted prevalences and in regression models. (see Supplemental Figure 2).

**Table 1:**
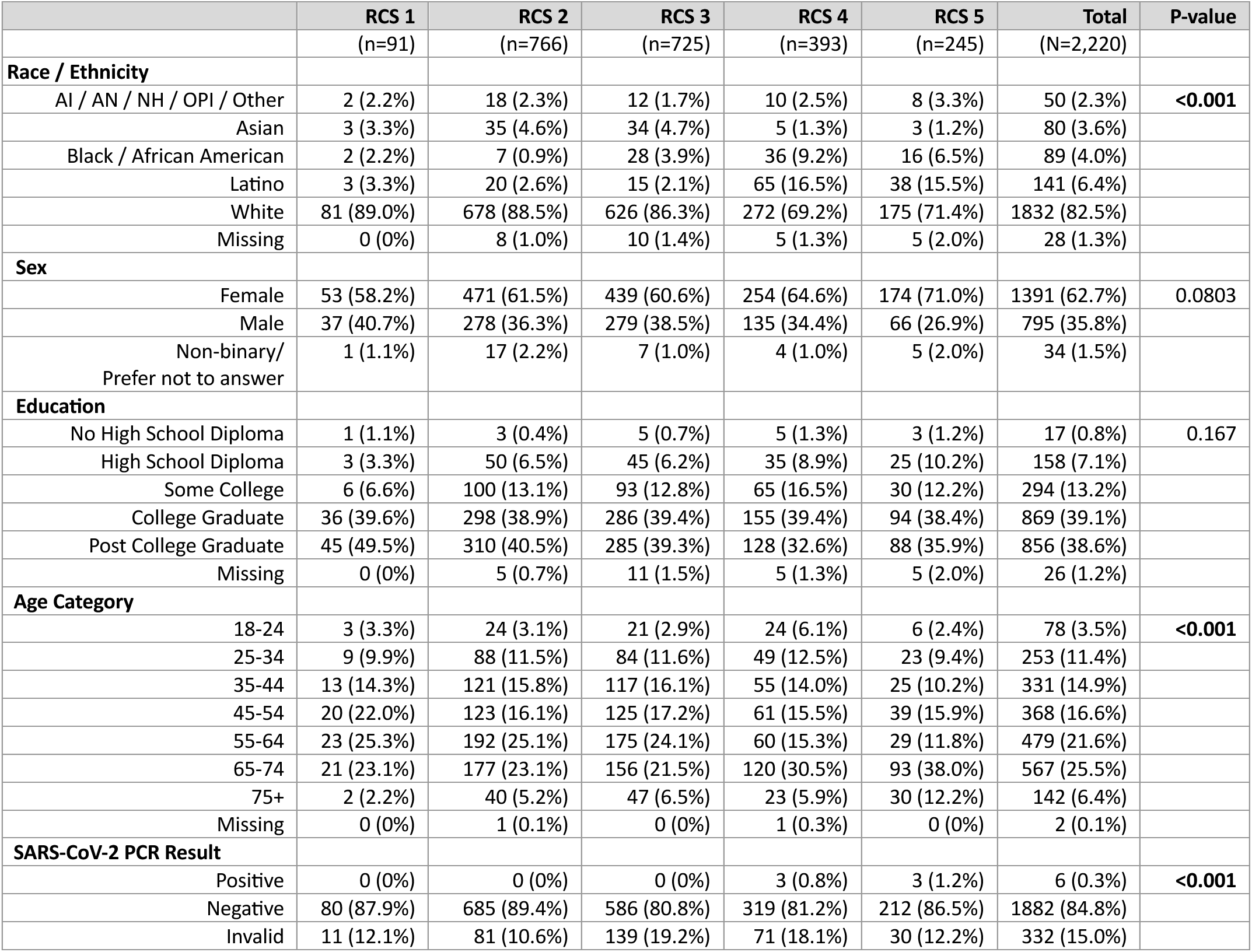
Basic Demographics of Study Participants in each RCS (unweighted), in the “Get Back Massachusetts” study, June 2021-September 2022. *Notes: AN = American Native; AI = American Indian; NH = Native Hawaiian; OPI = Other Pacific Islander* *Due to the limited number of participants in RCSs 1-2, these RCSs have been aggregated for survey weights and data analysis. ** Variables in boldface were used for IPF weighting.

### Population-level serostatus

Across all RCSs, the highest prevalence of seropositivity was found for spike antibodies. In the unweighted analysis, spike levels ranged from 86.4% of the population (RCS 3) to 97.4% of the population (RCS 5). After weighting, the results are broadly similar with a range of 83.1% to 96.4% seropositivity. Levels of the neutralizing antibody increased across each RCS with prevalence ranging from 20.5% (weighted, RCS 1 and 2) to 73.5% (weighted, RCS 5). Prevalence of capsid positivity remained relatively constant throughout the RCSs, except for a decrease to 32.9% in RCS 3 (Figure 2).

**Figure 2:**
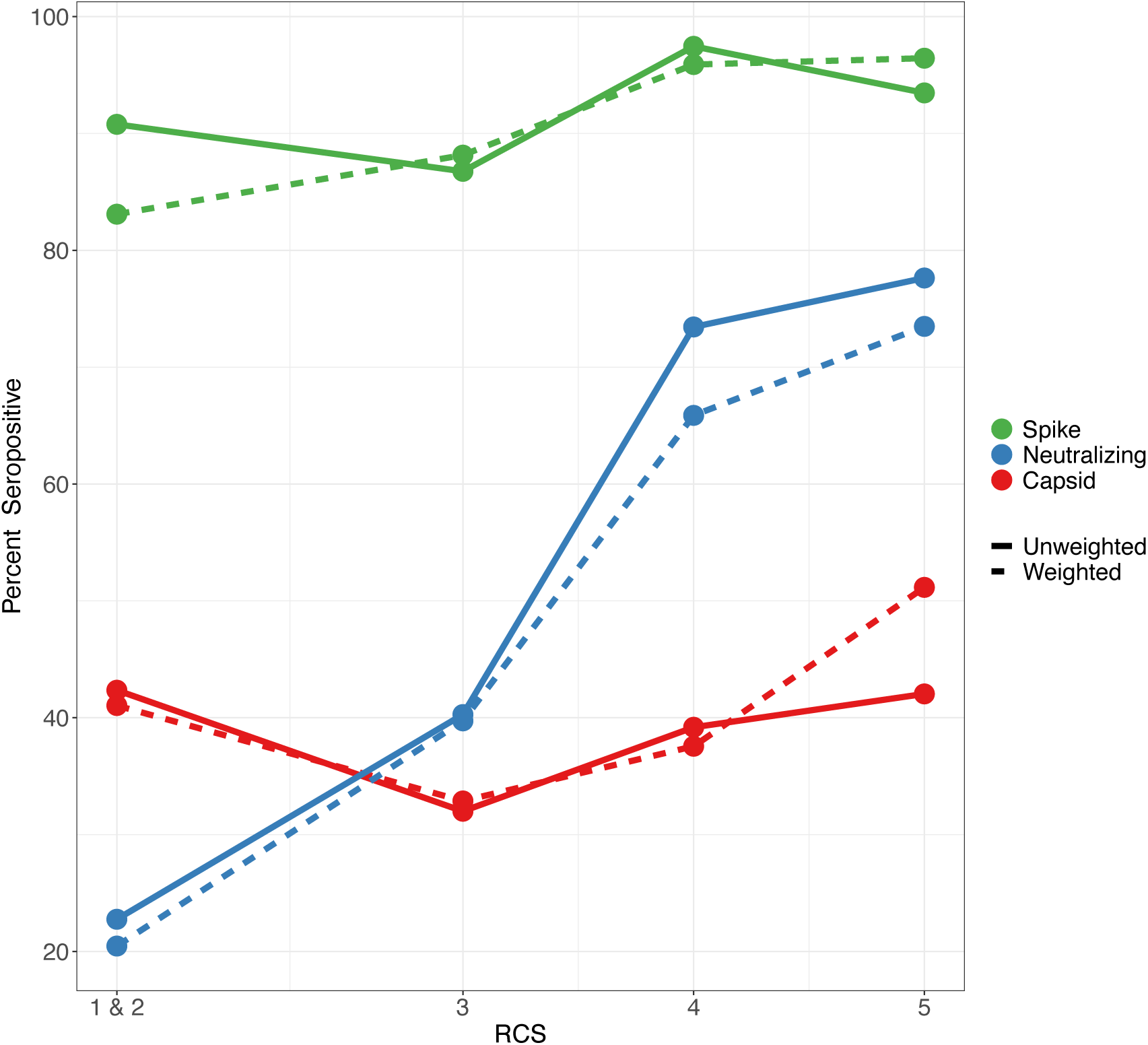
Proportion seropositive for antibodies to spike and capsid targets, and for spike neutralizing antibodies, by Repeated Cross-Sectional Survey (RCS), “Get Back Massachusetts” study, June 2021 - September 2022.

When stratified by vaccination status, differences emerge in neutralizing antibody seropositivity between vaccinated and unvaccinated participants. Unvaccinated individuals had lower weighted seroprevalence levels for all three markers, (capsid, neutralizing, and spike) compared to individuals who had been vaccinated. Importantly, only a small number of participants reported being unvaccinated. A stratified analysis highlights the very limited number of individuals who had not received any vaccination at the time of the blood sampling (Figure 3 and Supplemental Figure 5).

**Figure 3:**
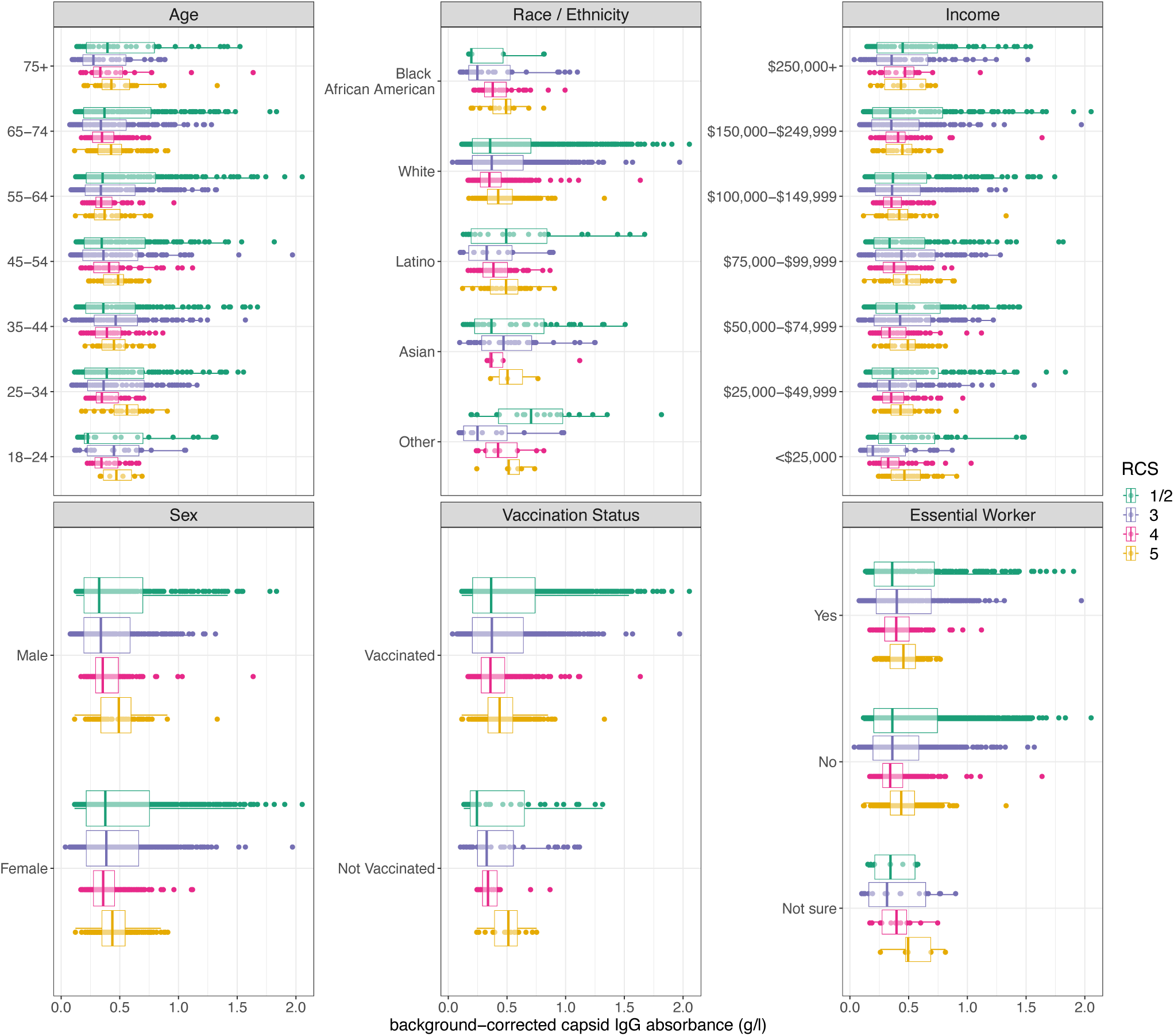
Corrected Capsid IgG antibody measures, by biodemographic characteristics across repeated cross-sectional surveys, “Get Back Massachusetts” study, June 2021 - September 2022. Notes: Boxplots show the median and interquartile ranges of the value for background-corrected capsid IgG absorbance for RCSs 1/2 (green), RCS 3 (purple), RCS 4 (pink), and RCS 5 (yellow). Data points within the boxplots show total number participants included for each category.

### Factors associated with capsid antibody serostatus

The distributions of measured capsid IgG titers, stratified by key demographic factors, and by RCS are shown in Figure 3. Overall, only modest differences can be observed. Within the age categories, the largest variation in median capsid IgG absorbance is observed among participants aged 18 – 24. In RCS 1/2, the median capsid IgG absorbance was 0.24 while in RCS 5, the median value increased to 0.47. For all other age groups, there were no major changes in median value (< 0.15). Assessing by Race/Ethnicity, there is a clear change in median population titer for the category “other”; the smallest median was observed in RCS 3 with the highest value during RCS 1-2. However, this group also had the fewest respondents (48 total). For “white” ethnicity, there was a large response (1,823), and comparisons of the median values show only minor variation between RCSs 1/2 and 5. There are no clear trends when median titer is compared across income groups. The median value for RCS 5 was 0.46, with a corresponding median of 0.19 at RCS; this difference is not statistically significant (via Kruskal-Wallis test). Additionally, there are no clear trends in median values when comparing by sex, or by essential worker status across the RCS rounds.

The Kruskal-Wallis test showed no statistically significant differences in median capsid IgG antibody titers between vaccinated and unvaccinated groups within each RCS round (p-values > 0.05 for all comparisons). Specifically, within RCS 1&2, RCS 3, RCS 4, and RCS 5, pairwise comparisons indicated only small, non-statistically significant differences between the vaccinated and unvaccinated groups. (Supplemental Figure 6).

In multivariable models to assess factors associated with capsid seropositivity (as a binary value), for RCS rounds 1/2 factors included in the final model were age, sex, race/ethnicity, income, wave (1 vs. 2), and vaccination status. The only variables that showed a statistically significant association with quantitative IgG antibody status (controlling for all other variables) were RCS and ethnicity (classified as “other”) (Table 2). Similarly, in multivariable models for RCS 5, the factors included in the final model were age, sex, race/ethnicity, income, and vaccination status. For the multivariable model for RCS 5, the factor found to be statistically significantly associated with quantitative IgG antibody status was race (African Americans and others (individuals who identify as American Native, American Indian, Native Hawaiian, or Other Pacific Islander). Specifically, Black/African American, and other persons had a higher seroprevalence for capsid (7.2% and 20.6% higher, respectively). Due to the limited numbers of participants in both ethnic classifications (Asian and AI / AN / NH / OPI / Other), confidence intervals surrounding these prevalence ratios are wide. There was no evidence for statistically significant differences in seroprevalence ratios by income categories (Table 2). Consistent results with only minor differences were obtained when modeling the titer values directly in multivariable quantile regressions, and can be found in the Supplemental Information, Table 2.

**Table 2:** Multivariable regression model results for seropositivity (SARS-CoV-2 capsid IgG) for RCS 1-2, and RCS 5 in the “Get Back Massachusetts” study, June 2021-September 2022. (Robust Poisson models to estimate prevalence ratios (PRs)).

|  | RCS 1 - 2 |  |  |  | RCS 5 |  |  |  |
| --- | --- | --- | --- | --- | --- | --- | --- | --- |
|  | PR | Lower<br>95% CI | Upper<br>95% CI | P-value | PR | Lower<br>95% CI | Upper<br>95% CI | p-value |
| **Age** | 1.002 | 0.998 | 1.005 | 0.399 | 0.999 | 0.995 | 1.002 | 0.423 |
| **Sex** |  |  |  |  |  |  |  |  |
| Female | Ref | - | - | - | Ref | - | - | - |
| Male | 0.902 | 0.805 | 1.012 | 0.078 | 1.085 | 0.962 | 1.224 | 0.186 |
| **Race / Ethnicity** |  |  |  |  |  |  |  |  |
| White | Ref | - | - | - | Ref | - | - | - |
| AI / AN / NH / OPI / Other | **1.485** | **1.098** | **2.009** | **0.01** | **1.206** | **1.000** | **1.455** | **0.050** |
| Asian | 1.175 | 0.922 | 1.497 | 0.193 | 1.051 | 0.978 | 1.129 | 0.174 |
| Black/African American | 0.667 | 0.389 | 1.143 | 0.141 | **1.072** | **1.009** | **1.139** | **0.025** |
| Latino | 1.046 | 0.721 | 1.518 | 0.814 | 1.032 | 0.932 | 1.143 | 0.543 |
| **Income** |  |  |  |  |  |  |  |  |
| < \$25,000 | Ref | - | - | - | Ref | - | - | - |
| \$25,000-49,999 | 0.991 | 0.742 | 1.323 | 0.951 | 0.936 | 0.8 | 1.095 | 0.407 |
| \$50,000-74,999 | 1.066 | 0.822 | 1.384 | 0.629 | 0.964 | 0.819 | 1.133 | 0.653 |
| \$75,000-99,999 | 0.933 | 0.718 | 1.212 | 0.604 | 1.023 | 0.851 | 1.229 | 0.811 |
| \$100,000-149,999 | 0.995 | 0.774 | 1.278 | 0.966 | 0.908 | 0.759 | 1.086 | 0.291 |
| \$150,000-249,999 | 1.027 | 0.787 | 1.339 | 0.846 | 0.925 | 0.777 | 1.101 | 0.378 |
| \$250,000+ | 1.07 | 0.819 | 1.397 | 0.622 | 0.949 | 0.712 | 1.265 | 0.722 |
| **Repeated Cross-sectional Survey** |  |  |  |  |  |  |  |  |
| RCS 1 | Ref | - | - | - | - | - | - | - |
| RCS 2 | 0.88 | 0.68 | 1.09 | 0.082 | - | - | - | - |
| RCS 5 | - | - | - | - |  |  |  |  |
| **Vaccination Status** |  |  |  |  |  |  |  |  |
| Not vaccinated | Ref | - | - | - | Ref | - | - | - |
| Vaccinated | 1.121 | 0.87 | 1.444 | 0.378 | 0.984 | 0.841 | 1.152 | 0.844 |

### Factors associated with neutralizing antibody status and distribution

An additional analysis of neutralizing antibodies evaluated changes over the RCS rounds (Supplemental Information Table 3). Two separate multivariable regressions were undertaken to compare factors associated with neutralizing antibody seropositivity between early and later RCS rounds.

The first model contained data collected during RCSs 1 and 2; the second model contained data collected in RCS 5. These models include only the subset of respondents who were seropositive for capsid. (n= 781 for RCS 1/2; and n= 228 for RCS 5). In the first model, the only variables with a statistically significant association with neutralizing antibody status (controlling for age, sex, race/ethnicity, and income) were RCS round (1 vs 2), and vaccination status. In the corresponding model for RCS 5, vacation status, race, and income were all found to be statistically significantly associated with neutralizing antibody status after controlling for age and sex (Supplemental Information Table 4).

Finally, we evaluated the aggregate prevalence of neutralizing antibodies by county in Massachusetts during RCSs 1 and 2, and then in RCS 5. During the first RCSs of this survey, antibody prevalences ranged from 8.7% in Essex to 37.8% prevalence in Franklin County. During the final wave of the survey, there was an increase in reported prevalence across the state. The largest increase in prevalence occurred in Barnstable County (10.7% to 92.2%). During RCS 5, the highest recorded prevalence was in Franklin County in which 100% of participants tested positive for neutralizing antibodies. (Supplemental Information Figure 3).

## IV. Discussion and Conclusions

This study found that there were no major differences in antibody titers were observed across RCS or by demographics (other than vaccination status) indicating near-universal pathogen exposure and suggesting that all population groups were similarly protected from SARS-CoV-2 infection statewide, with no major disparities among vulnerable populations. This is an important finding as it suggests that across diverse demographic groups, there were no sub-populations that had unusually high levels of antibodies relative to other demographics. While these data cannot disentangle the joint impacts of vaccine-induced antibodies from those that are subsequent to infection, it strongly suggests no populations in the state had excessive prior infection or unusually high vaccination relative to other groups (23).

Across the five RCSs, there was a progressive increase in detectable neutralizing antibodies, ranging from 22.8% in RCS 1 to 77.6% in RCS 5, particularly spiking from 40.3% in RCS 3 to 73.5% in RCS 4, coinciding with the Omicron-1 variant outbreak and heightened vaccination uptake in Massachusetts. This surge aligns with rollout of the mRNA-based COVID-19 vaccines (Pfizer-BioNTech and Moderna) throughout the state, which produces antibodies to the viral spike protein (24,25). In contrast, detectable capsid antibodies were at moderate levels (32.0% - 42.4%) across the five RCSs, potentially due to a “steady state” of waning capsid antibodies approximately six months post-infection, and the fact that most COVID-19 vaccines target the spike protein rather than the nucleocapsid protein (25,26). Spike positivity remained consistently high (over 90%) across RCSs, suggesting that a majority of participants had had exposure to viral antigens, either through primary infection or vaccination.

Considering capsid response (infection-related), an additional finding is the inverse association observed between the prevalence ratios of capsid positivity and vaccination status across the RCSs. This unexpected result may be attributed to several factors, including small sample sizes in the unvaccinated group, or a potential residual confounding factor associated with both contracting SARS-CoV-2 and receiving the vaccine. It is also plausible that individuals who did not receive the vaccine had prior SARS-CoV-2 exposure earlier in the pandemic, and there might be some waning antibody levels among this group. However, these findings are not definitive and require further investigation.

These seroprevalence estimates are a combination of two trends-seroconversion and seroreversion rates, and represent a “snapshot” of these two competing processes. A recent systematic review reports high sensitivity for detection of RBD/spike antibodies up to 6 months, while nucleocapsid titers wane more rapidly, potentially having sensitivity as low as 57% at 6 months post-exposure (27). Although our study results cannot directly assess the impact of seroreversion on estimated seroprevalence, it provides valuable insights into antibody dynamics during the Omicron wave (Dec 2021 to Feb 2022). Finally, this study is unable to estimate population-level immunity from neutralizing antibody measurements, as thresholds for determining potential protection from infection (“correlates of protection”) are still being refined (28,29).

In comparison with other COVID-19 reporting (clinical testing, hospitalization, and mortality data) serology surveys are a critical tool to track transmission, as they can better capture sub-clinical or asymptomatic infection and historical infection. In this study, we highlight the results of the largest to our knowledge population-based serosurvey of seroprevalence among Massachusetts residents. Conducted over sixteen months (June 2021 – September 2022) and across eight different SARS-CoV-2 variants, this set of cross-sectional studies allows for a better understanding of COVID-19 seroprevalence across the state at different times, in different subpopulations, and using different seromarkers of SARS-CoV-2 exposure.

This study’s strengths include the large sample size; a rigorous state-level sample frame; detailed socio-demographic information; biological sample collection from at-home testing kits; and a repeated survey design to provide insights into viral exposures over several waves of the pandemic. Another strength is that compared to other work in Massachusetts measuring SARS-CoV-2 seroprevalence, this study is notably larger and encompasses a broader population, contrasting with the more targeted approach of the Boston (Mass General) study which concentrated on healthcare workers (30). Limitations include a less diverse population due to limited response rates, an inability to track individual titer responses over time as participants were included at a single time point, and uncertainty regarding the generalizability of results to future SARS-CoV-2 variants.

Future studies should consider longitudinal sampling of individuals to better measure changes in antibody titers in respond to factors like co-infections, vaccinations, and repeated exposures. Despite directed oversampling of underrepresented population groups, we observed higher response rates for women, Caucasians, people in high-income households, and highly educated people, prompting the need for post-sampling IPF raking to enhance representativeness; future studies should explore alternative survey strategies to more effectively recruit individuals with lower education levels and from underrepresented racial groups. Lastly, given the high vaccination coverage in Massachusetts, future research should oversample unvaccinated individuals to investigate differences between vaccinated and unvaccinated populations.

## Data Availability

All data produced in the present study are available upon reasonable request to the authors

## Supplemental figures and tables

**Supplemental Figure 1:**
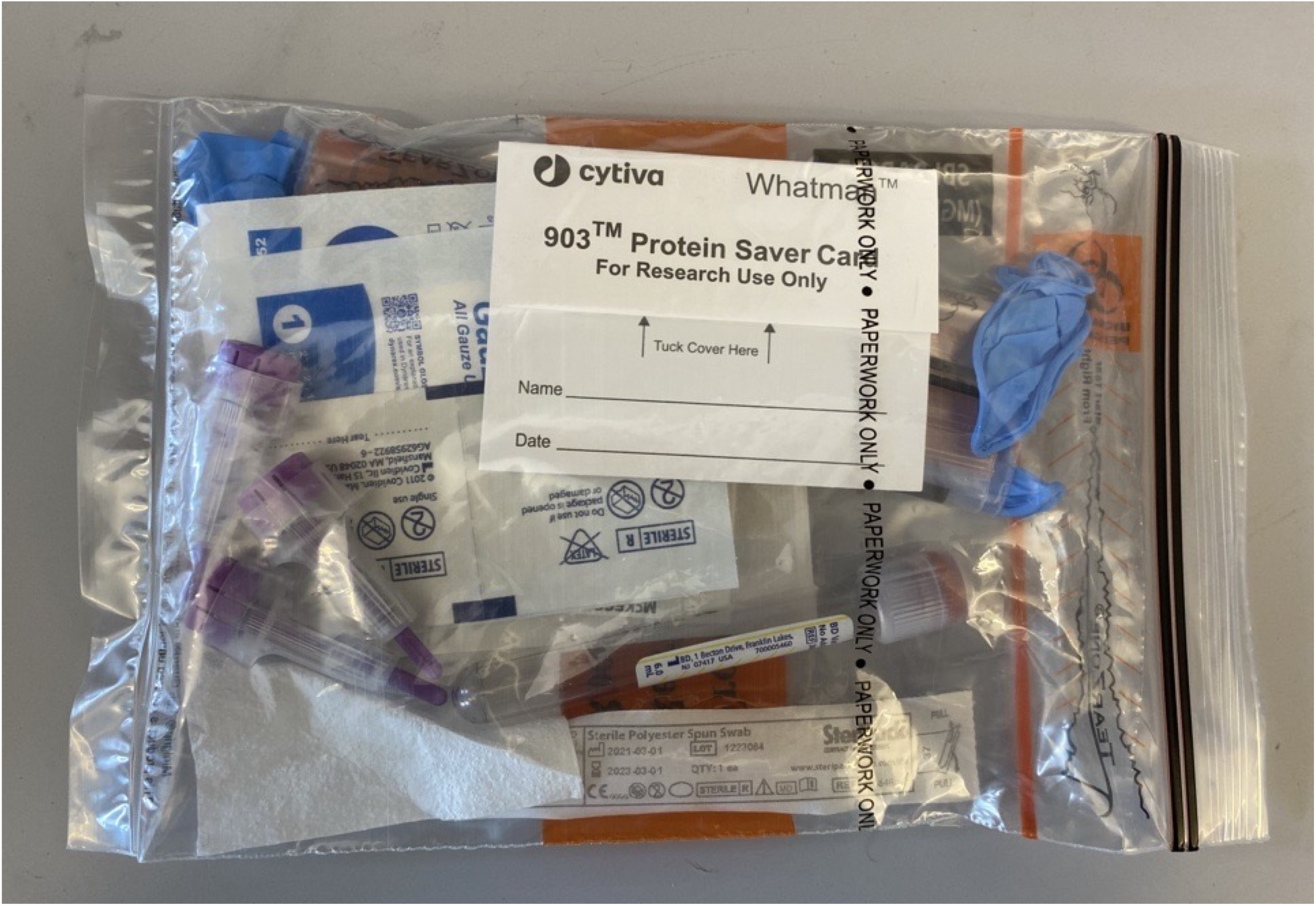
Biosample sample collection kit, “Get Back Massachusetts” study, June 2021-September 2022.

**Supplemental Figure 2:**
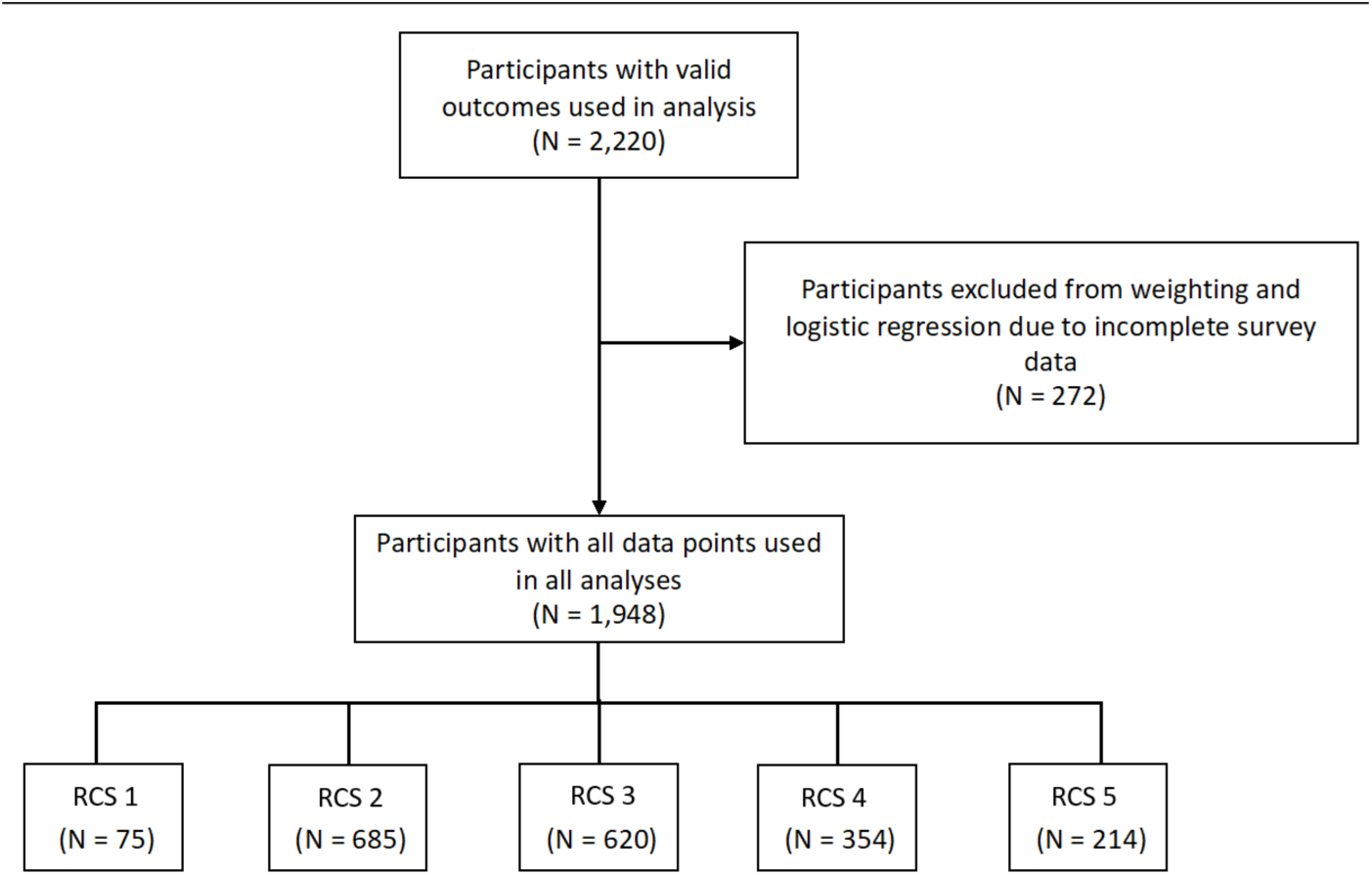
Participants excluded in IPF raking and logistic regression due to missing data, “Get Back Massachusetts” study, June 2021-September 2022.

**Supplemental Figure 3:**
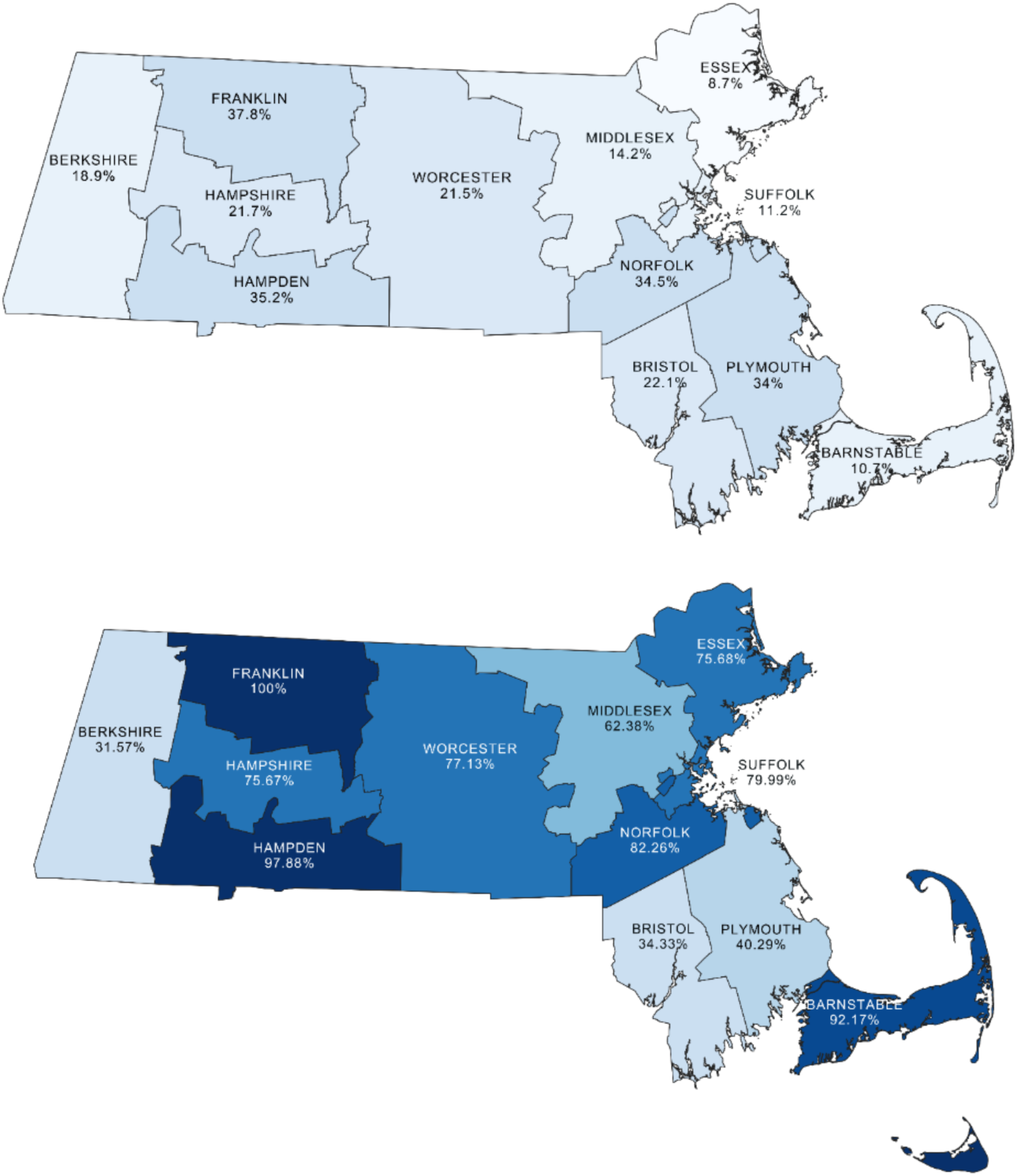
Neutralizing Antibody weighted and stratified by county, “Get Back Massachusetts” study, June 2021-September 2022. Notes. Panel A shows positivity calculated during RCSs 1-2; panel B shows seropositivity during RCS 5. Prevalence of seropositivity was calculated using weighted values

**Supplemental Table 1:** Spike, neutralizing, and capsid, positive by RCS round of the Covid-19 pandemic in the “Get Back Massachusetts” study, June 2021-September 2022. Notes: *IPF raking utilized categorical variables, *Age*, *Race*, *Sex*, and *Education*. For participant weighting, participants with missing data were excluded from this analysis.

| RCS | Proportion Spike Positive |  | Proportion with Detectable Neutralizing antibodies |  | Proportion with Detectable Capsid antibodies |  |
| --- | --- | --- | --- | --- | --- | --- |
|  | Unweighted | Weighted | Unweighted | Weighted | Unweighted | Weighted |
| 1 and 2 | 90.8% | 83.1% | 22.8% | 20.5% | 42.4% | 41.0% |
| 3 | 86.8% | 88.1% | 40.3% | 39.7% | 32.0% | 32.9% |
| 4 | 97.5% | 95.9% | 73.4% | 65.9% | 39.2% | 37.6% |
| 5 | 93.5% | 96.4% | 77.6% | 73.5% | 42.0% | 51.2% |

**Supplemental Table 2:**
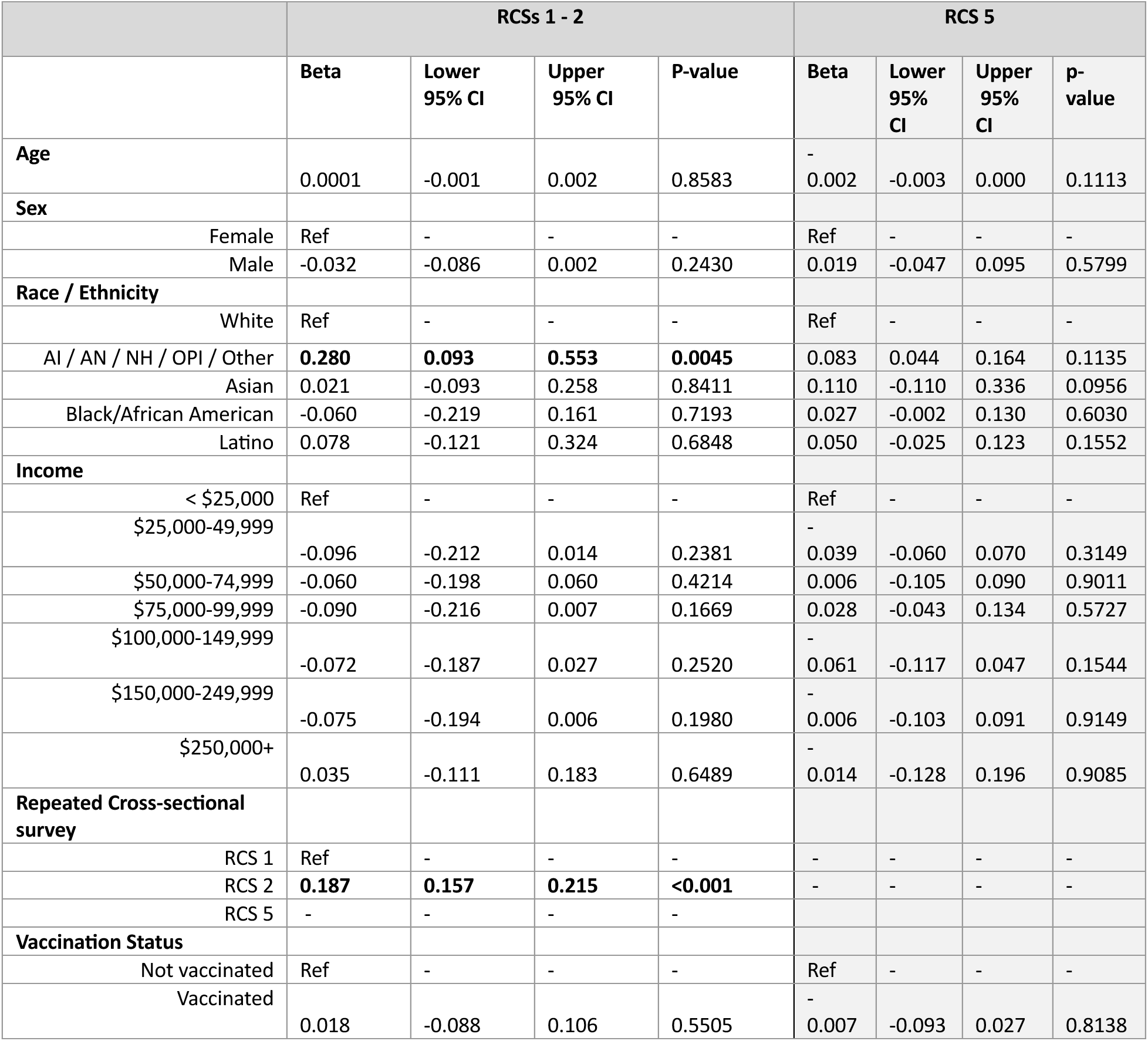
Multivariable quantile (median) regression model for quantified IgG capsid antibodies to SARS-CoV-2 for RCS 1-2, and RCS 5 in the “Get Back Massachusetts” study, June 2021-September 2022). (Quantile regression models to estimate relationships between capsid antibody titers and covariates).

**Supplemental Table 3:**
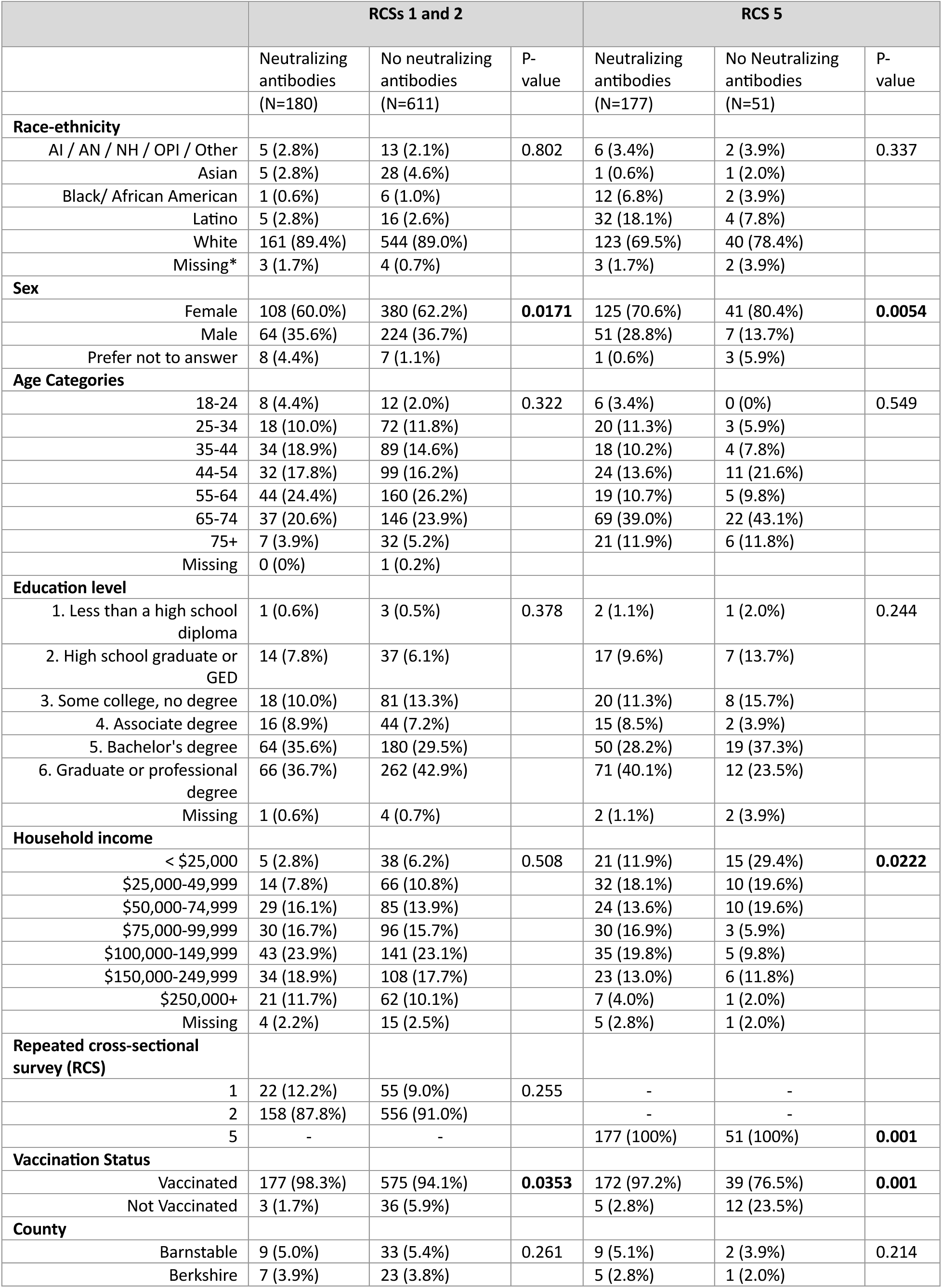
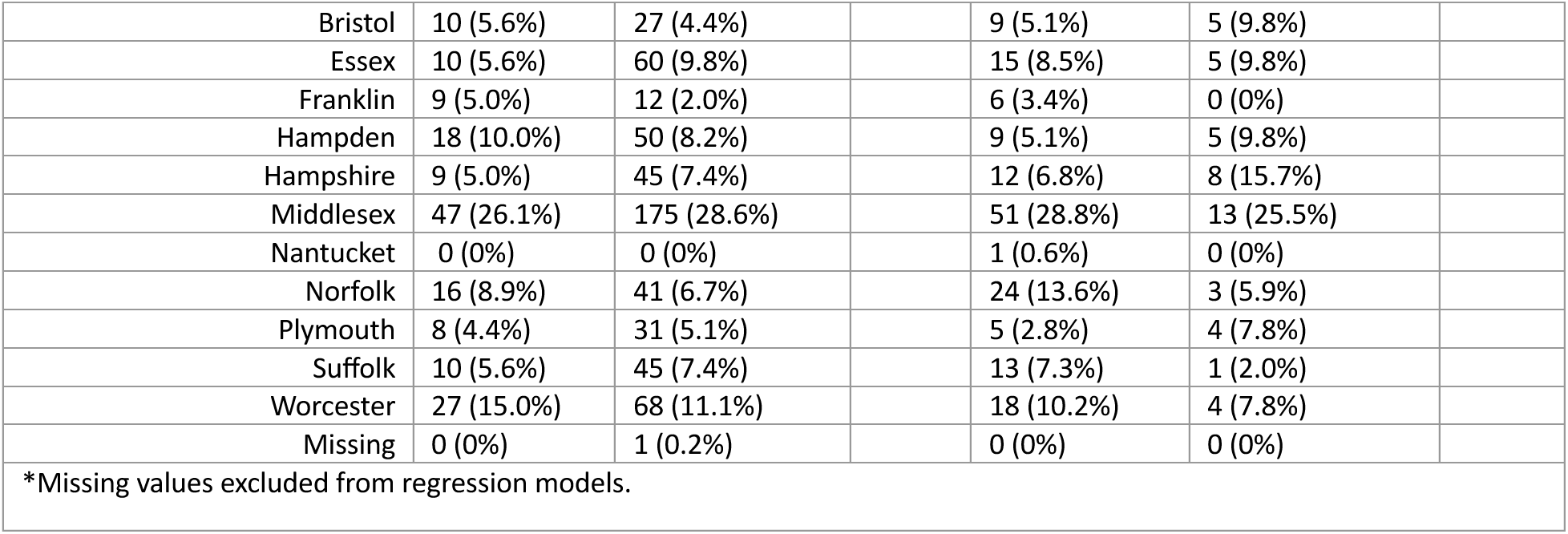
Comparison of demographic factors associated with seropositivity for neutralizing antibodies: RCSs 1 and 2 (aggregated) versus RCS 5 in the “Get Back Massachusetts” study, June 2021-September 2022. Note: p-value for difference from χ^2^ tests.

|  | RCSs 1 and 2 |  |  | RCS 5 |  |  |
| --- | --- | --- | --- | --- | --- | --- |
|  | Neutralizing antibodies | No neutralizing antibodies | P-value | Neutralizing antibodies | No Neutralizing antibodies | P-value |
|  | (N=180) | (N=611) |  | (N=177) | (N=51) |  |
| **Race-ethnicity** |  |  |  |  |  |  |
| AI / AN / NH / OPI / Other | 5 (2.8%) | 13 (2.1%) | 0.802 | 6 (3.4%) | 2 (3.9%) | 0.337 |
| Asian | 5 (2.8%) | 28 (4.6%) |  | 1 (0.6%) | 1 (2.0%) |  |
| Black/ African American | 1 (0.6%) | 6 (1.0%) |  | 12 (6.8%) | 2 (3.9%) |  |
| Latino | 5 (2.8%) | 16 (2.6%) |  | 32 (18.1%) | 4 (7.8%) |  |
| White | 161 (89.4%) | 544 (89.0%) |  | 123 (69.5%) | 40 (78.4%) |  |
| Missing* | 3 (1.7%) | 4 (0.7%) |  | 3 (1.7%) | 2 (3.9%) |  |
| **Sex** |  |  |  |  |  |  |
| Female | 108 (60.0%) | 380 (62.2%) | 0.0171 | 125 (70.6%) | 41 (80.4%) | 0.0054 |
| Male | 64 (35.6%) | 224 (36.7%) |  | 51 (28.8%) | 7 (13.7%) |  |
| Prefer not to answer | 8 (4.4%) | 7 (1.1%) |  | 1 (0.6%) | 3 (5.9%) |  |
| **Age Categories** |  |  |  |  |  |  |
| 18-24 | 8 (4.4%) | 12 (2.0%) | 0.322 | 6 (3.4%) | 0 (0%) | 0.549 |
| 25-34 | 18 (10.0%) | 72 (11.8%) |  | 20 (11.3%) | 3 (5.9%) |  |
| 35-44 | 34 (18.9%) | 89 (14.6%) |  | 18 (10.2%) | 4 (7.8%) |  |
| 44-54 | 32 (17.8%) | 99 (16.2%) |  | 24 (13.6%) | 11 (21.6%) |  |
| 55-64 | 44 (24.4%) | 160 (26.2%) |  | 19 (10.7%) | 5 (9.8%) |  |
| 65-74 | 37 (20.6%) | 146 (23.9%) |  | 69 (39.0%) | 22 (43.1%) |  |
| 75+ | 7 (3.9%) | 32 (5.2%) |  | 21 (11.9%) | 6 (11.8%) |  |
| Missing | 0 (0%) | 1 (0.2%) |  |  |  |  |
| **Education level** |  |  |  |  |  |  |
| 1. Less than a high school diploma | 1 (0.6%) | 3 (0.5%) | 0.378 | 2 (1.1%) | 1 (2.0%) | 0.244 |
| 2. High school graduate or GED | 14 (7.8%) | 37 (6.1%) |  | 17 (9.6%) | 7 (13.7%) |  |
| 3. Some college, no degree | 18 (10.0%) | 81 (13.3%) |  | 20 (11.3%) | 8 (15.7%) |  |
| 4. Associate degree | 16 (8.9%) | 44 (7.2%) |  | 15 (8.5%) | 2 (3.9%) |  |
| 5. Bachelor's degree | 64 (35.6%) | 180 (29.5%) |  | 50 (28.2%) | 19 (37.3%) |  |
| 6. Graduate or professional degree | 66 (36.7%) | 262 (42.9%) |  | 71 (40.1%) | 12 (23.5%) |  |
| Missing | 1 (0.6%) | 4 (0.7%) |  | 2 (1.1%) | 2 (3.9%) |  |
| **Household income** |  |  |  |  |  |  |
| < \$25,000 | 5 (2.8%) | 38 (6.2%) | 0.508 | 21 (11.9%) | 15 (29.4%) | 0.0222 |
| \$25,000-49,999 | 14 (7.8%) | 66 (10.8%) | | 32 (18.1%) | 10 (19.6%) | |
| \$50,000-74,999 | 29 (16.1%) | 85 (13.9%) | | 24 (13.6%) | 10 (19.6%) | |
| \$75,000-99,999 | 30 (16.7%) | 96 (15.7%) | | 30 (16.9%) | 3 (5.9%) | |
| \$100,000-149,999 | 43 (23.9%) | 141 (23.1%) | | 35 (19.8%) | 5 (9.8%) | |
| \$150,000-249,999 | 34 (18.9%) | 108 (17.7%) | | 23 (13.0%) | 6 (11.8%) | |
| \$250,000+ | 21 (11.7%) | 62 (10.1%) | | 7 (4.0%) | 1 (2.0%) | |
| Missing | 4 (2.2%) | 15 (2.5%) |  | 5 (2.8%) | 1 (2.0%) |  |
| **Repeated cross-sectional survey (RCS)** |  |  |  |  |  |  |
| 1 | 22 (12.2%) | 55 (9.0%) | 0.255 | - | - |  |
| 2 | 158 (87.8%) | 556 (91.0%) |  | - | - |  |
| 5 | - | - |  | 177 (100%) | 51 (100%) | 0.001 |
| **Vaccination Status** |  |  |  |  |  |  |
| Vaccinated | 177 (98.3%) | 575 (94.1%) | 0.0353 | 172 (97.2%) | 39 (76.5%) | 0.001 |
| Not Vaccinated | 3 (1.7%) | 36 (5.9%) |  | 5 (2.8%) | 12 (23.5%) |  |
| **County** |  |  |  |  |  |  |
| Barnstable | 9 (5.0%) | 33 (5.4%) | 0.261 | 9 (5.1%) | 2 (3.9%) | 0.214 |
| Berkshire | 7 (3.9%) | 23 (3.8%) |  | 5 (2.8%) | 1 (2.0%) |  |
| Bristol | 10 (5.6%) | 27 (4.4%) |  | 9 (5.1%) | 5 (9.8%) |  |
| Essex | 10 (5.6%) | 60 (9.8%) |  | 15 (8.5%) | 5 (9.8%) |  |
| Franklin | 9 (5.0%) | 12 (2.0%) |  | 6 (3.4%) | 0 (0%) |  |
| Hampden | 18 (10.0%) | 50 (8.2%) |  | 9 (5.1%) | 5 (9.8%) |  |
| Hampshire | 9 (5.0%) | 45 (7.4%) |  | 12 (6.8%) | 8 (15.7%) |  |
| Middlesex | 47 (26.1%) | 175 (28.6%) |  | 51 (28.8%) | 13 (25.5%) |  |
| Nantucket | 0 (0%) | 0 (0%) |  | 1 (0.6%) | 0 (0%) |  |
| Norfolk | 16 (8.9%) | 41 (6.7%) |  | 24 (13.6%) | 3 (5.9%) |  |
| Plymouth | 8 (4.4%) | 31 (5.1%) |  | 5 (2.8%) | 4 (7.8%) |  |
| Suffolk | 10 (5.6%) | 45 (7.4%) |  | 13 (7.3%) | 1 (2.0%) |  |
| Worcester | 27 (15.0%) | 68 (11.1%) |  | 18 (10.2%) | 4 (7.8%) |  |
| Missing | 0 (0%) | 1 (0.2%) |  | 0 (0%) | 0 (0%) |  |
| *Missing values excluded from regression models. |  |  |  |  |  |  |

**Supplemental Table 4:** Multivariable model results for seroprevalence of neutralizing antibodies to SARS-CoV-2 for RCS 1-2, and RCS 5 in the “Get Back Massachusetts” study, June 2021-September 2022). Note: (Robust Poisson models to estimate prevalence ratios (PRs).

|  | RCSs 1 - 2 |  |  |  | RCS 5 |  |  |  |
| --- | --- | --- | --- | --- | --- | --- | --- | --- |
|  | PR | Lower 95% CI | Upper 95% CI | P-value | PR | Lower 95% CI | Upper 95% CI | p-value |
| **Age** | 0.99 | 0.98 | 1.00 | 0.075 | 0.99 | 0.99 | 1.00 | 0.79 |
| **Sex** |  |  |  |  |  |  |  |  |
| Female | Ref |  |  |  | Ref |  |  |  |
| Male | 0.97 | 0.74 | 1.29 | 0.86 | 1.10 | 0.97 | 1.26 | 0.15 |
| **Race / Ethnicity** |  |  |  |  |  |  |  |  |
| White | Ref |  |  |  | Ref |  |  |  |
| AI / AN / NH / OPI / Other | 1.22 | 0.54 | 2.76 | 0.63 | 1.13 | 0.73 | 1.72 | 0.54 |
| Asian | 0.59 | 0.24 | 1.48 | 0.26 | 0.66 | 0.14 | 3.15 | 0.60 |
| Black/African American | 0.73 | 0.12 | 4.47 | 0.74 | **1.20** | **0.99** | **1.45** | **0.044** |
| Latino | 0.9 | 0.36 | 2.24 | 0.82 | **1.22** | **1.01** | **1.42** | **0.010** |
| **Income** |  |  |  |  |  |  |  |  |
| < \$25,000 | Ref | | | | Ref | | | |
| \$25,000-49,999 | 1.34 | 0.51 | 3.45 | 0.58 | 1.19 | 0.87 | 1.62 | 0.27 |
| \$50,000-74,999 | 1.87 | 0.76 | 4.70 | 0.15 | 1.08 | 0.77 | 1.51 | 0.65 |
| \$75,000-99,999 | 1.80 | 0.73 | 4.42 | 0.18 | **1.42** | **1.07** | **1.88** | **0.014** |
| \$100,000-149,999 | 1.72 | 0.72 | 4.26 | 0.19 | **1.39** | **1.03** | **1.87** | **0.031** |
| \$150,000-249,999 | 1.76 | 0.73 | 4.55 | 0.15 | 1.30 | 0.95 | 1.76 | 0.11 |
| \$250,000+ | 1.77 | 0.70 | 4.67 | 0.18 | 1.32 | 0.91 | 1.91 | 0.15 |
| **Repeated Cross-Sectional Survey** |  |  |  |  |  |  |  |  |
| RCS 1 | Ref | - | - | - | - | - | - | - |
| RCS 2 | **0.67** | **0.46** | **0.99** | **0.042** | - | - | - | - |
| RCS 5 | - | - | - | - | Ref |  |  |  |
| **Vaccination Status** |  |  |  |  |  |  |  |  |
| Not vaccinated | Ref | - | - | - | Ref | - | - | - |
| Vaccinated | **2.88** | **1.00** | **8.24** | **0.049** | **2.91** | **1.32** | **6.41** | **0.008** |

**Supplemental Figure 4:**
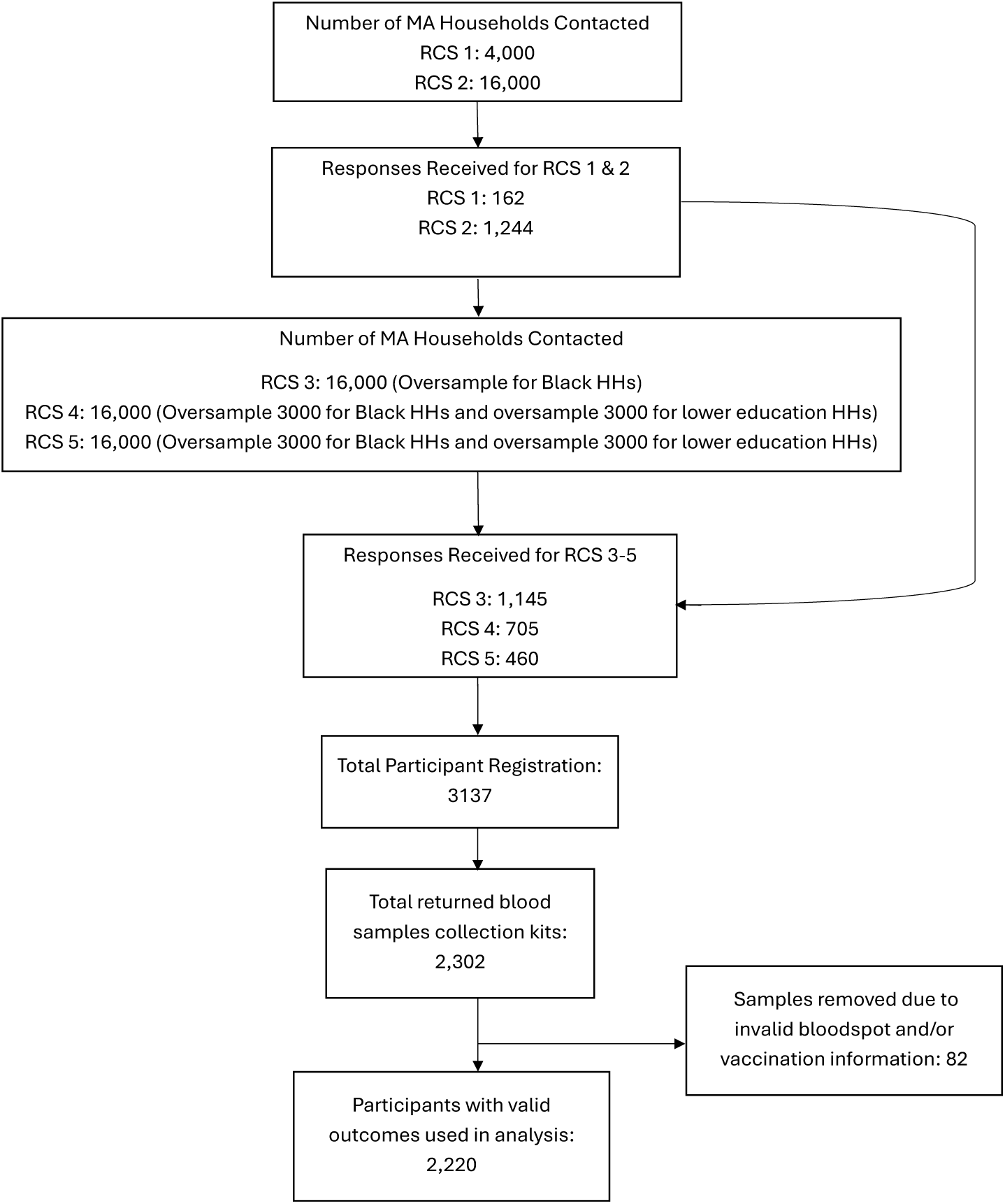
Study enrollment (CONSORT) diagram, “Get Back Massachusetts” study, June 2021 - September 2022.

**Supplemental Figure 5:**
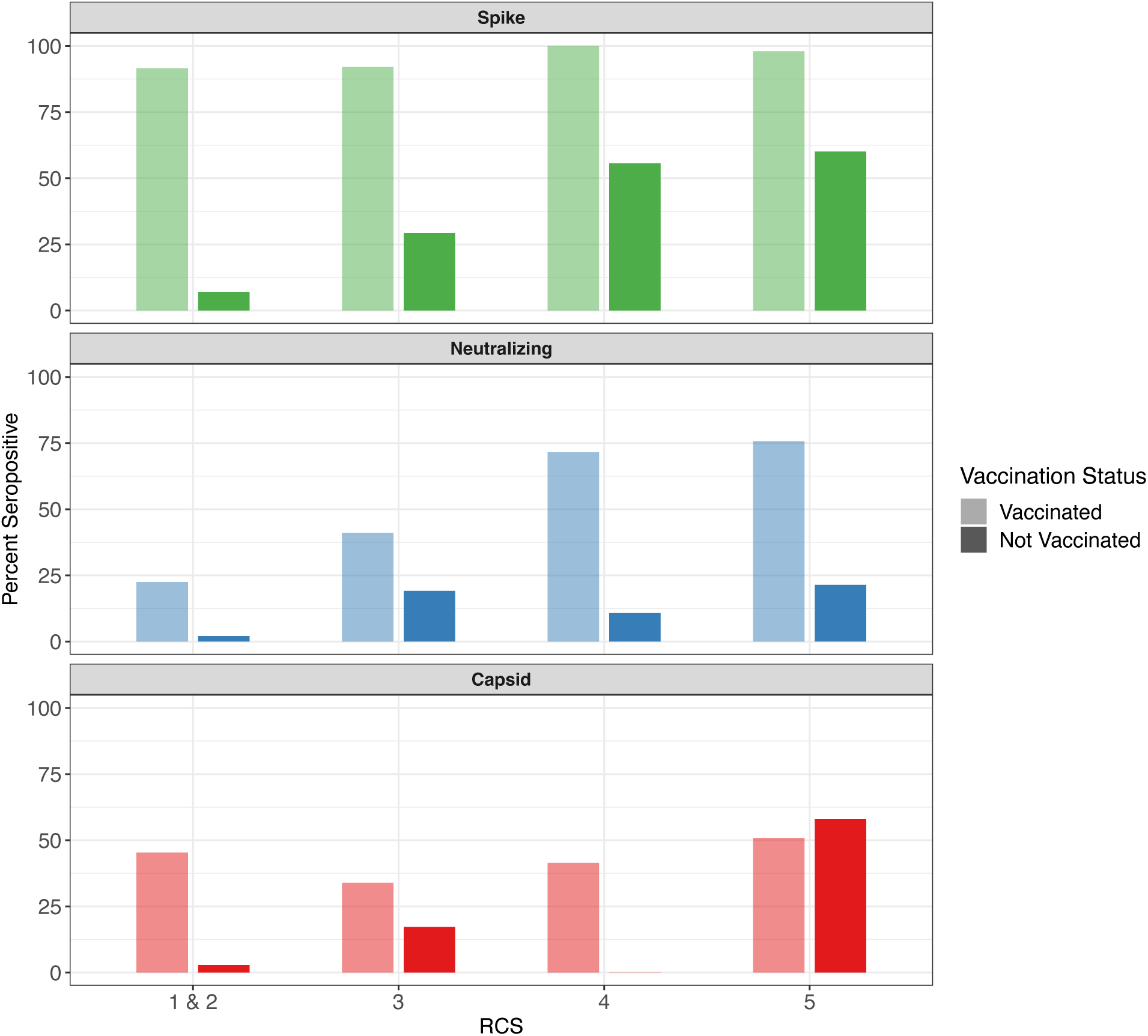
Prevalence of capsid, neutralizing, and spike by Repeated Cross-Sectional Survey (RCS) stratified by vaccine status, “Get Back Massachusetts” study, June 2021-September 2022.

**Supplemental Figure 6:**
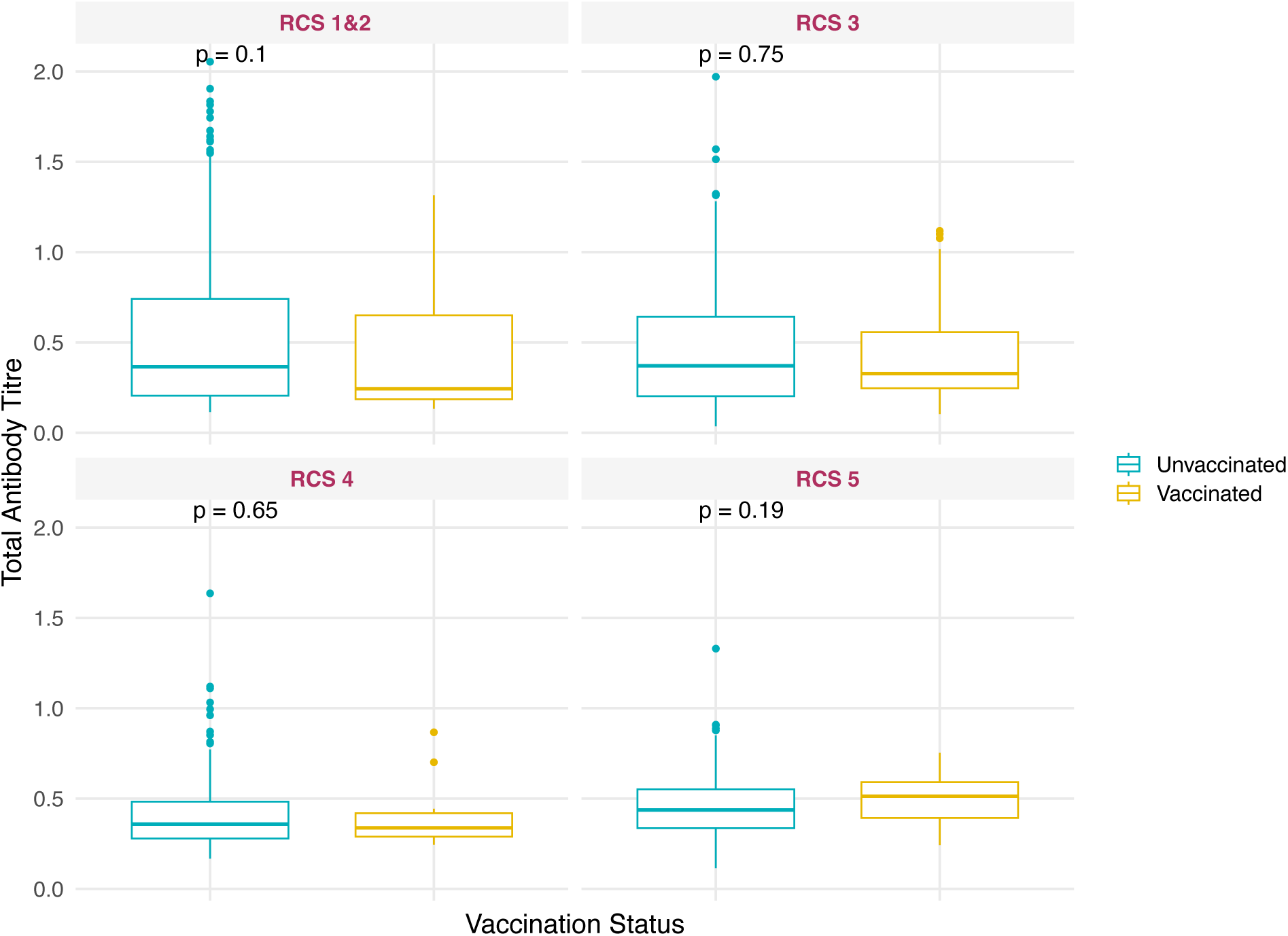
Comparisons of median capsid IgG antibody titers by vaccination status across RCS, “Get Back Massachusetts” study, June 2021-September 2022. (p-value from individual Kruskal-Wallis tests).

## Supplemental File 1

### Details on laboratory analyses and neutralizing antibody model development

**Neutralizing**: To categorize the output for the neutralizing antibody measurements, output from the neutralizing antibody test was converted to relative Light Unit (RLU). RLU readings were converted to a percent neutralization efficiency (PNE) value using the following equation:

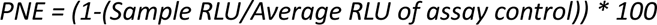

**Capsid**: The presence of human antibodies bound to the viral antigen was determined using a secondary anti-human immunoglobulin antibody conjugated to horseradish peroxidase. Chromogenic substrate addition and absorption measurement at threshold (OD_405_ = 0.468 OR 0.426) identified samples with antibodies reactive to SARS CoV-2 capsid protein.

**Neutralizing antibody model development**: To determine whether there was variation in factors related to neutralizing antibodies (a combination of infection and vaccine-induced exposures) changed over the RCSs of data collection, two separate multivariable regressions were performed using complete case data. The first model analyzed RCSs 1/2 together (earliest data) and the second model analyzed data from RCS 5 (latest data). The outcome for both models was the likelihood of neutralizing antibody seropositivity. This approach allowed for a more nuanced understanding of the dynamics and determinants of antibody response over the course of RCSs 1 - 5, potentially providing valuable insights into the evolving nature of immune responses to the antigen. Demographic characteristics by binary neutralizing antibody serostatus are shown in Supplemental Table 3.

The variables for RCSs 1/2 that were included in the final model were age, sex, race/ethnicity, income, wave (1 vs. 2), and vaccination status. The only variables with a statistically significant association with neutralizing antibody status (controlling for all other variables) were RCS and vaccination status. Prevalence ratios are shown in Supplemental Table 4. Similarly, multivariable models for RCS 5, the factors included in the final model were age, sex, race/ethnicity, income, and vaccination status. In the multivariable model for RCS 5, the factors found to have statistically significantly associations with neutralizing antibody status were vaccination status, race, and income. Specifically, vaccinated persons had 2.9-fold (95% CI: 1.3 to 6.4-fold) times the seroprevalence of unvaccinated persons. Also, relative to white respondents, Black/African American and Latino persons had higher risk of seroprevalence for neutralizing antibodies (21% and 22% higher, respectively). Asian individuals had a lower risk of neutralizing antibody prevalence compared to white respondents (PR = 0.66, 95% CI: 0.73 to 1.72); individuals who identify as American Native, American Indian, Native Hawaiian, Other Pacific Islander, or any other unlisted race have 1.13 times the likelihood of testing positive for neutralizing antibodies (95% CI: 0.73-1.72). Due to the low numbers of participants in both ethnic classifications (Asian and AI / AN / NH / OPI / Other), there is much uncertainty surrounding the prevalence ratios reported. Finally, relative to lowest income households, in RCS 5, residents of households reporting incomes of $75,000-99,000 and $100,000-150,000 had higher prevalence of neutralizing antibodies (42% and 39% higher, respectively than households reporting an income below $25,000. With the ordinal classification of income, there is a bi-directional trend in prevalence ratios across income groups; the highest prevalence occurs for the income category $75,000 - $99,999 and the lowest prevalence occurs for the income category $50,000-74,999 (PR = 1.08) (Supplemental Table 4).

